# Operational Survival Deficit of Neoadjuvant Chemotherapy in Early-Stage Breast Cancer: A Target Trial Emulation and Causal Machine Learning Study

**DOI:** 10.64898/2025.12.22.25342768

**Authors:** Shihao Guan, Yangyang Jian, Wanying Dong, Liangpeng Dong

## Abstract

**Background:** Neoadjuvant chemotherapy (NAC) is the standard of care for locally advanced breast cancer. However, the disconnect between efficacy in randomized trials and effectiveness in real-world practice—attributable to real-world treatment delays and adherence barriers—remains underexplored for early-stage (cT1-cT3) operable disease.

**Methods:** We applied the Target Trial Emulation (TTE) framework to a propensity-score matched cohort from the SEER database. To mitigate immortal time bias and staging migration, we reconstructed clinical baselines. Individualized Treatment Effects (ITE) were estimated using a Double-Robust Causal Forest algorithm. To rigorously cross-validate these estimates against model misspecification, we employed a DeepCox neural network as a non-linear sensitivity analysis tool, exposing complex risk structures (e.g., U-shaped hazards) that traditional linear assumptions might overlook.

**Results:** In the matched cohort (N=26,946), Standard NAC was associated with an operational survival deficit (Absolute Risk Difference: 3.6%) compared to upfront surgery, corresponding to a hazard ratio of 1.32 (95% CI, 1.24–1.40; *p* < 0.001). Causal Forest analysis revealed a critical “Response-Survival Discordance”: while young TNBC patients exhibited high nodal pathologic complete response (npCR) rates, they paradoxically faced the worst survival outcomes (Standard Cox HR 1.87). Even in the 6-month landmark analysis to account for immortal time bias, this survival detriment persisted (Landmark HR 1.39; 95% CI, 1.06–1.81; *p* = 0.016; Figure 3D). Crucially, node-positive (cN+) patients—traditionally considered ideal candidates for systemic downstaging—experienced a significant survival detriment with NAC (HR 1.39). This disadvantage was most pronounced in Luminal A subtype and Invasive Lobular Carcinoma (ILC), where NAC failed to provide effective source control. In contrast, HER2-positive status exhibited a trend towards survival benefit, diverging from the significant risks observed in other subtypes. Anatomically, while cT2 tumors identified a “window of minimal operational deficit” where the absolute risk difference was negligible, operational risk paradoxically resurged in cT3 tumors, challenging the conventional paradigm that larger burdens inherently mandate downstaging.

**Conclusion:** Our causal analysis reveals a critical disconnect between biological risk and therapeutic efficacy. While SHAP modeling identified node-positive (cN+) status as a high-priority indicator for systemic therapy, the low real-world response rate (npCR 15.0%) rendered historical standard NAC regimens insufficient to counterbalance the risks of surgical delay (HR 1.39). Our findings indicate that without therapeutic escalation (e.g., immunotherapy) to ensure high pathologic response rates, the operational risks of deferring surgery may outweigh the benefits of downstaging in this subgroup. Our findings highlight a critical “Implementation Gap” where standard NAC regimens yield suboptimal real-world outcomes for high-risk subgroups. Our findings suggest that clinical prioritization should diverge based on subtype biology: for chemo-refractory subtypes (e.g., Luminal A, ILC), Upfront Surgery ensures immediate source control and should be prioritized; conversely, for high-risk TNBC, standard NAC is insufficient, warranting Therapeutic Escalation (e.g., immunotherapy) to minimize the risk of non-response.

## 1. Introduction

Breast cancer remains the leading cause of cancerrelated morbidity and mortality among women worldwide [6, 34]. While incidence peaks in postmenopausal women, the diagnosis in young women (<40 years) is rising and is characterized by historically less favorable prognosis compared with older cohorts [36]. Young age (≤ 40 years) is an independent predictor of adverse outcomes (HR 1.4) [28]. While international guidelines (BCY5) recommend NAC for this demographic to facilitate breast conservation [10, 27], our study challenges whether this recommendation accounts for the rapid progression risks in real-world settings. Neoadjuvant chemotherapy (NAC) has evolved from a downstaging tool for locally advanced disease to a standard-of-care recommendation for early-stage (cT1-cT3) Triple-Negative Breast Cancer (TNBC) [15, 21]. Current guidelines increasingly favor NAC to utilize pathologic complete response (pCR) as an *in vivo* chemosensitivity test [9, 25, 37] to guide adjuvant escalation [33].

However, a critical dissonance persists between *clinical trial efficacy* and *real-world effectiveness*. While landmark RCTs (e.g., NSABP B-18) established the **biological efficacy** of NAC under idealized conditions [4, 14], they do not account for the **“Systemic Friction”** of routine practice— defined here as the cumulative hazard of treatment delays, toxicity-induced interruptions, and variable adherence [31]. Unlike the idealized conditions of RCTs, this study evaluates the **programmatic effectiveness** of the neoadjuvant *strategy*, capturing the **“Total Treatment Hazard”**—which encompasses not only pharmacological efficacy but also the operational risks of treatment intervals and adherence variations [30].

For young patients with rapidly proliferating TNBC, the 4-6 month preoperative window is not merely a monitoring period but a potential window of disease progression where treatment delays can be detrimental [1, 5, 7, 16] For young patients with TNBC, this creates a critical tension between the theoretical benefit of drug sensitivity testing and the immediate need for **“Time to definitive local therapy.”** Biologically, these tumors are characterized by rapid doubling times and high proliferation indices [24, 32]. For this demographic, the 4-6 month preoperative chemotherapy window is not merely a monitoring period but a potential window for micrometastatic progression if the tumor proves chemo-refractory [3, 19]. In the presence of adherence barriers, the delay in definitive surgical control may outweigh the benefits of downstaging.

Therefore, the critical question arises: *In the imperfect reality of clinical practice, for which specific subgroups does the potential for pCR justify the risk of delaying definitive surgery?* To answer this, we adopted a **multilayered methodological framework**: (1) **Target Trial Emulation (TTE)** and **Propensity Score Matching (PSM)** to enforce strict comparability analogous to randomized trials; (2) **Causal Forest algorithms** to unmask heterogeneous treatment effects often hidden in standard regression; and (3) **Deep Learning sensitivity analyses** (DeepCox) to validate non-linear risk topographies. This rigorous triangulation aims to isolate biological signals from potential statistical artifacts.

## 2. Methods

### 2.1. Data Source and Study Population

To emulate a hypothetical target trial using observational data, we adhered to the framework described by Hernán and Robins [17]. This approach aligns the start of follow-up with the treatment decision point, thereby minimizing prevalent user bias and immortal time bias.

Data were retrieved from the Surveillance, Epidemiology, and End Results (SEER) 18 Registries database (November 2020 submission), covering approximately 28% of the U.S. population [26]. Institutional Review Board (IRB) approval was waived given the de-identified, publicly available nature of the data. We utilized the 1975– 2018 dataset to leverage Collaborative Stage (CS) variables, which are essential for precise tumor sizing and extension characterization [12].

We identified female patients diagnosed with primary invasive breast cancer between 2010 and 2015. This timeframe was selected to ensure a minimum of 5-year follow-up while reflecting contemporary treatment standards. The inclusion criteria were strictly defined as follows: (1) Unilateral, first primary malignancy; (2) Histologically confirmed Invasive Ductal Carcinoma (IDC) or Invasive Lobular Carcinoma (ILC); and (3) Clinically operable disease (cT1-cT3, N0-N+, M0).

To ensure a homogeneous “clean cohort” for causal inference, we applied rigorous exclusion criteria: (1) Distant metastasis (M1) at diagnosis; (2) T4 disease, including inflammatory breast cancer or direct extension to the chest wall/skin, as these cases lack clinical equipoise for upfront surgery; (3) Unknown treatment sequencing or missing biological markers (Grade, ER, PR, HER2 status), to prevent bias from imputation.

### 2.2. Construction of the Clinical Restaging System

To eliminate the “Time-Travel Bias” pervasive in previous registry studies, we constructed a rigorous **Clinical Restaging System** based on raw tumor size and physical extension codes, reconstructing the pre-treatment baseline independent of pathological downstaging.

Unlike standard analyses that rely on derived TNM stages (often confounded by post-treatment pathology, ypT), our approach creates a strictly “clean” pre-treatment baseline comparable to RCT enrollment [17].

We utilized raw *Collaborative Stage (CS)* variables rather than derived stages. Specifically:

- **Tumor Extension:** *CS Extension* codes 400–800 (involving skin/chest wall invasion) were strictly mapped to cT4 and excluded to ensure clinical equipoise.
- **Tumor Size:** *CS Tumor Size* code 990 (Microscopic) was imputed to 0.5mm, ensuring precise continuous variable measurement.
- **Nodal Status:** Clinical N stages (cN) were reconstructed using *CS Lymph Nodes* codes to differentiate clinically node-negative (cN0) from node-positive (cN+) disease, independent of pathological findings.

This logic-based reconstruction ensures that the treatment assignment (NAC vs. Upfront Surgery) is evaluated based on the information available *at diagnosis*, effectively preventing immortal time bias.

### 2.3. Variable Definitions and Endpoints

Patients were stratified into two intention-to-treat strategies based on the temporal sequence of therapy: the **NAC Group** (systemic therapy initiated before surgery) and the **Upfront Surgery Group** (surgery performed as the initial therapeutic intervention).

The primary endpoint was Breast Cancer-Specific Survival (BCSS), defined as the time from diagnosis to death attributable to breast cancer. The secondary endpoint was Overall Survival (OS). As an exploratory biological endpoint, we evaluated the nodal pathologic Complete Response (npCR) rate within the NAC cohort, defined as the conversion from cN+ to ypN0.

### 2.4. Statistical Analysis: The Double-Robust Framework

To address the inherent selection bias in observational data, we employed a “Double-Robust” inference strategy combining propensity score matching with machine learning estimation [18].

#### 2.4.1. Propensity Score Matching and Cohort Construction

We calculated the propensity score (PS) for receiving NAC using a multivariable logistic regression model. Co-variates were strictly limited to pre-treatment baseline characteristics, including age, race, marital status, income, ruralurban residence, year of diagnosis, histology, grade, molecular subtype, and reconstructed clinical T/N stages. Post-treatment variables (e.g., surgery type) were explicitly excluded from the propensity score estimation to preclude selection bias. To ensure comparability, we performed “common support trimming” by excluding upfront surgery patients whose PS fell below the 1st percentile of the NAC distribution. We then executed 1:1 nearest-neighbor matching with a strict caliper of 0.02. Covariate balance was assessed using the Standardized Mean Difference (SMD), with a threshold of < 0.1 indicating negligible imbalance (visualized in Figure S3).

#### 2.4.2. Causal Inference Strategy and Heterogeneity Analysis

Beyond average treatment effects, we utilized the **Causal Forest** algorithm with **orthogonalization** to isolate the treatment effect from confounders [35]. To prevent overfitting, we implemented **sample splitting (“honesty”)**: one half of the data was used to build the tree structure, and the other half to estimate the treatment effects. This non-parametric approach allows for the detection of complex, non-linear treatment effect heterogeneity across subgroups (e.g., young TNBC or elderly HER2+ patients) without pre-specifying interaction terms. To ensure model interpretability and identify specific drivers of heterogeneity (e.g., Age), we further employed SHAP (Shapley Additive exPlanations) values utilizing the TreeExplainer algorithm [23]. **To mitigate variance underestimation inherent in ensemble methods, 95% confidence intervals (CIs) for the Average Treatment Effect (ATE) were derived using 1**,**000 bootstrap iterations**.

#### 2.4.3. Sensitivity Assessment: Capturing Non-Linear Risk Structures

Standard Cox proportional hazards models rely on linear assumptions, which may fail to detect complex biological patterns such as threshold effects. To test the robustness of our causal findings against potential *linear model misspecification*, we utilized a **DeepCox** neural network [20]. In this study, DeepCox serves not as a primary clinical prediction tool, but as a **validation instrument** to uncover non-linear risk topographies (e.g., U-shaped hazards) and high-order interactions. Model discrimination (C-index) and calibration were assessed specifically to confirm that the identified non-linear heterogeneity is structurally real and not an artifact of algorithmic overfitting (performance metrics detailed in Figure S1 and Figure S2).

### 2.5. Sensitivity Analysis

To rule out “immortal time bias” (where NAC patients must survive long enough to complete chemotherapy and reach surgery), we conducted a **Landmark Analysis**, excluding all patients who died or were censored within 6 months of diagnosis. Additionally, we calculated the **E-value** to quantify the strength of unmeasured confounding (e.g., Ki-67 index) required to explain away the observed treatment effects. **To rigorously assess the stability of our findings against the temporal evolution of chemotherapy regimens, we performed a Time-Interaction Analysis, testing the interaction between treatment strategy and year of diagnosis**.

Furthermore, to ensure **Model-Agnostic Robustness**, we validated our findings using three additional metalearning algorithms: **S-Learner, T-Learner, and X-Learner** [22], benchmarking them against the Causal Forest estimates (Figure 2E, F). All analyses were performed using Python (v3.10) with *lifelines, econml*, and *pytorch* libraries [11, 29]. Statistical significance was set at two-sided *p* < 0.05. The reporting of this study follows the TRIPOD+AI statement and the Strengthening the Reporting of Observational Studies in Epidemiology (STROBE) reporting guideline [8].

**Figure 1:**
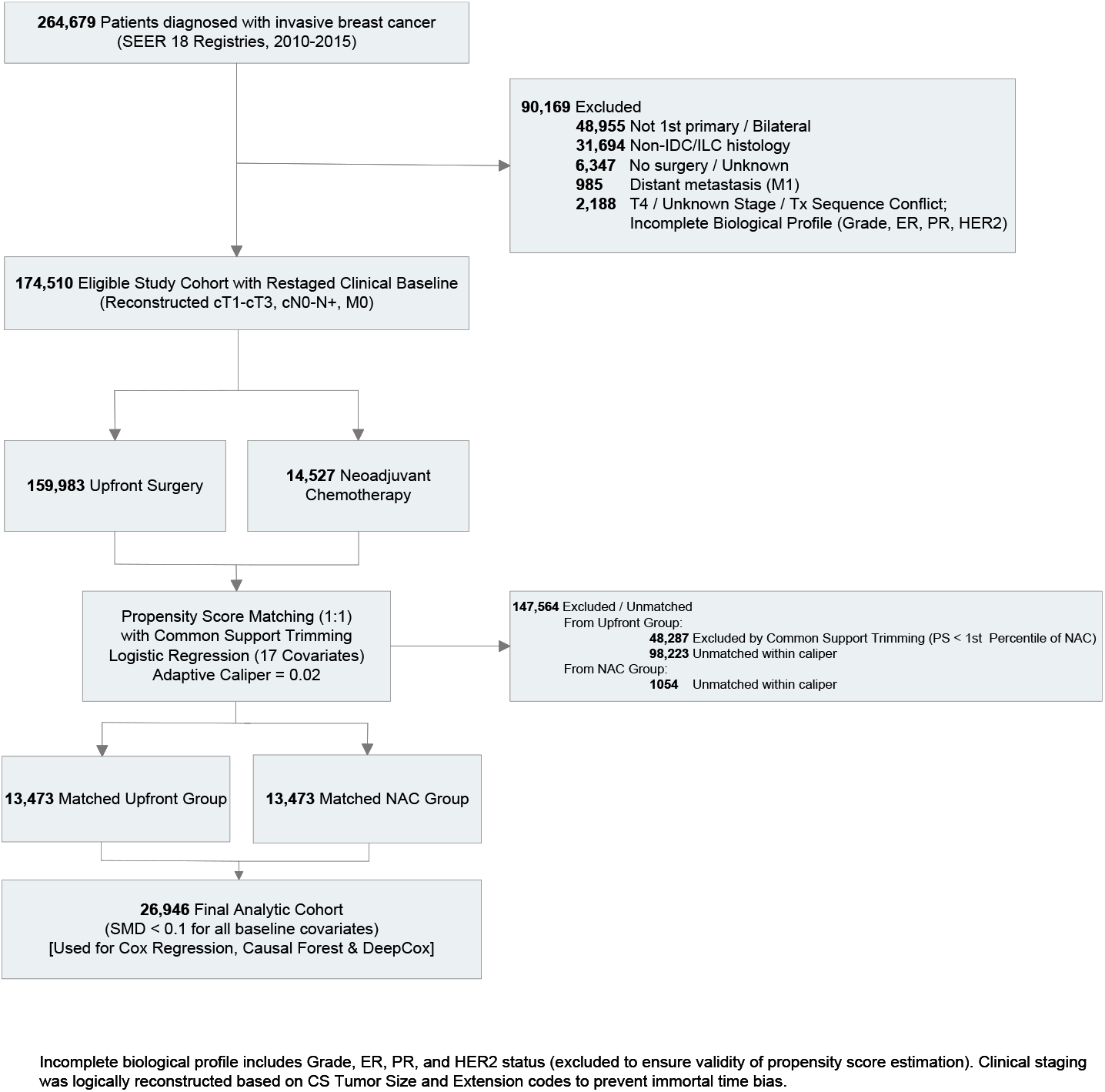
Study Flowchart. A total of 264,679 patients were initially identified. After applying rigorous exclusion criteria—including the use of a **Clinical Restaging System** to eliminate **staging migration bias**—and conducting 1:1 Propensity Score Matching, 26,946 patients were included in the final analysis.

**Figure 2:**
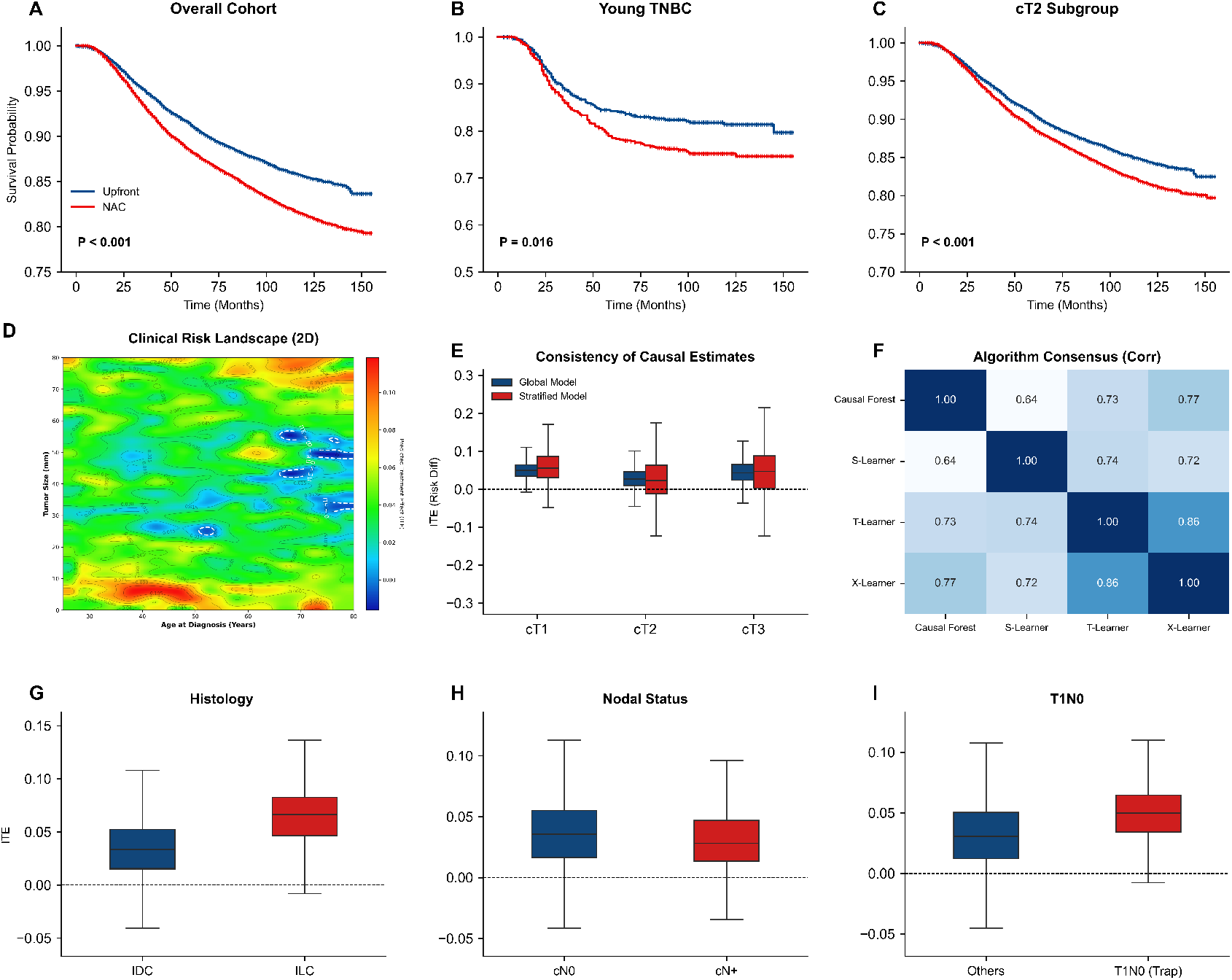
Multidimensional Analysis of Treatment Effect Heterogeneity and Subgroup Outcomes. Row 1 (Survival Outcomes) Kaplan-Meier estimates of overall survival. (A) The overall matched cohort shows a statistically significant survival difference favoring upfront surgery. (B) The young TNBC subgroup (*<*40 years) demonstrates a pronounced survival disadvantage with NAC (6-Month Landmark Analysis). (C) The cT2 subgroup exhibits closely aligned survival curves, indicating that the magnitude of survival operational deficit is least pronounced in this anatomical window. **Row 2 (Model Validation):** (D) Heatmap of Individualized Treatment Effects (ITE) predicted by DeepCox, illustrating the distribution of risk across continuous variables. (E) **Consistency Analysis:** Stratified causal estimates by T-stage align with the global model, confirming algorithmic stability. (F) Correlation matrix among meta-learners validating the consensus of causal estimation. **Row 3 (Subgroup Analysis):** (G) **Histological Subtypes:** Invasive Lobular Carcinoma (ILC) is associated with a higher magnitude of NAC-related survival compromise compared to Invasive Ductal Carcinoma (IDC). (H) **Nodal Status:** Patients with clinically node-positive (cN+) disease exhibit a greater hazard ratio for mortality under NAC compared to the cN0 subgroup. (I) **Early-Stage Disease:** The T1N0 subgroup demonstrates a substantial survival disadvantage, suggesting disproportionate risks of NAC in early-stage, node-negative patients.

## 3. Results

### 3.1. Cohort Characteristics and Propensity Score Matching

The initial query of the SEER database yielded 264,679 patients (Figure 1). Following the rigorous exclusion of M1 disease, T4 stage, and cases with incomplete biological data, the final eligible study cohort was identified. Before matching, the NAC group presented with significantly higher risk features (e.g., higher grade, larger tumor size) compared to the Upfront Surgery group.

After implementing 1:1 propensity score matching with a strict caliper of 0.02, a balanced cohort of **26**,**946 patients** was retained (13,473 in each group). Baseline characteristics of the unmatched cohort are detailed in Table 1. The matching process achieved exceptional covariate balance, with the Standardized Mean Difference (SMD) for all baseline covariates falling well below 0.1, creating a quasi-randomized analytical framework (Figure S3).

**Table 1.** Baseline Characteristics of the Study Population Before Propensity Score Matching.

| Characteristic | Category | Unmatched Cohort (N=174,510) |  | SMD | P Value |
| --- | --- | --- | --- | --- | --- |
|  |  | NAC (N=14,527) | Upfront (N=159,983) |  |  |
| **Age (years)** | Mean (SD) | 52.0 (11.8) | 61.2 (12.9) | 0.723 | <0.001 |
| **Tumor Size (mm)** | Mean (SD) | 38.6 (48.1) | 24.2 (74.9) | 0.197 | <0.001 |
| **Year of Diagnosis** | Mean (SD) | 2012.9 (1.7) | 2012.6 (1.7) | 0.191 | <0.001 |
| **Household Income (US\$)** | Mean (SD) | 79,923 (18,909) | 80,401 (19,533) | 0.025 | 0.004 |
| **Race/Ethnicity** |  |  |  | 0.181 | <0.001 |
| White | n (%) | 10,641 (73.2%) | 128,210 (80.1%) |  |  |
| Black | n (%) | 2,210 (15.2%) | 14,887 (9.3%) |  |  |
| Asian/PI | n (%) | 1,501 (10.3%) | 15,205 (9.5%) |  |  |
| Other/Unknown | n (%) | 175 (1.2%) | 1,681 (1.1%) |  |  |
| **Marital Status** |  |  |  | 0.247 | <0.001 |
| Married | n (%) | 8,403 (57.8%) | 92,130 (57.6%) |  |  |
| Unmarried/Other | n (%) | 6,124 (42.2%) | 67,853 (42.4%) |  |  |
| **Rural/Urban** |  |  |  | 0.097 | <0.001 |
| Urban | n (%) | 13,362 (92.0%) | 143,211 (89.5%) |  |  |
| Rural | n (%) | 1,151 (7.9%) | 16,554 (10.3%) |  |  |
| **Clinical T Stage** |  |  |  | 1.028 | <0.001 |
| cT1 (<20mm) | n (%) | 3,322 (22.9%) | 109,454 (68.4%) |  |  |
| cT2 (20-50mm) | n (%) | 8,348 (57.5%) | 45,367 (28.4%) |  |  |
| cT3 (>50mm) | n (%) | 2,857 (19.7%) | 5,162 (3.2%) |  |  |
| **Clinical N Stage** |  |  |  | 0.656 | <0.001 |
| cN0 (Node Negative) | n (%) | 6,270 (43.2%) | 118,163 (73.9%) |  |  |
| cN+ (Node Positive) | n (%) | 8,257 (56.8%) | 41,820 (26.1%) |  |  |
| **Molecular Subtype** |  |  |  | 0.808 | <0.001 |
| TNBC (HR-/HER2-) | n (%) | 3,656 (25.2%) | 15,397 (9.6%) |  |  |
| HER2+ (All) | n (%) | 4,970 (34.2%) | 20,666 (12.9%) |  |  |
| Luminal A (HR+/HER2-) | n (%) | 5,901 (40.6%) | 123,920 (77.5%) |  |  |
| **Histologic Type** |  |  |  | 0.172 | <0.001 |
| Ductal (IDC) | n (%) | 13,677 (94.1%) | 143,114 (89.5%) |  |  |
| Lobular (ILC) | n (%) | 850 (5.9%) | 16,869 (10.5%) |  |  |
| **Histologic Grade** |  |  |  | 0.636 | <0.001 |
| Grade I-II (Well/Mod.) | n (%) | 5,883 (40.5%) | 113,090 (70.7%) |  |  |
| Grade III (Poorly Diff.) | n (%) | 8,644 (59.5%) | 46,893 (29.3%) |  |  |
| **Surgery Type** |  |  |  | 0.331 | <0.001 |
| Breast Conserving Surgery | n (%) | 5,653 (38.9%) | 99,840 (62.4%) |  |  |
| Mastectomy | n (%) | 8,874 (61.1%) | 60,143 (37.6%) |  |  |
| **Radiation Therapy** |  |  |  | 0.237 | <0.001 |
| Yes | n (%) | 9,257 (63.7%) | 89,433 (55.9%) |  |  |
| No/Unknown | n (%) | 5,270 (36.3%) | 70,550 (44.1%) |  |  |

Note: SMD, Standardized Mean Difference. High SMDs (>0.1) indicate significant baseline imbalances, necessitating propensity score matching. Surgery Type is a post-treatment variable presented for descriptive purposes.

### 3.2. Overall Treatment Effect and Mortality Risk

In the matched cohort, the Causal Forest analysis estimated a **modest but consistent absolute risk increase** in breast cancer-specific mortality (Mean Absolute Risk Difference ≈ 3.6%) for patients undergoing NAC compared to Upfront Surgery. Consistent with this absolute difference, the Cox proportional hazards model demonstrated a statistically significant relative risk increase (**HR 1.32**; 95% CI, 1.24–1.40; *p*< 0.001; Figure 2A).

However, the distribution of Individualized Treatment Effects (ITE) revealed a complex risk landscape (Figure 3A, B). Survival benefit from NAC was confined to a **highly specific, biomarker-defined subset** (predominantly HER2+). In contrast, the analysis identified a substantial **“At-Risk Cohort”**—comprising **over 40% of the population**—that exhibited a statistically significant **preference for Upfront Surgery** (ITE favoring surgery). This skewed distribution indicates that for a large segment of unselected patients, the operational risks of delay outweigh the potential benefits of downstaging, underscoring the necessity for precise, biomarker-driven patient selection. Furthermore, to capture risk patterns beyond linear assumptions, we utilized restricted cubic splines to map the non-linear relationship between tumor burden and mortality risk (Figure 4A).

**Figure 3:**
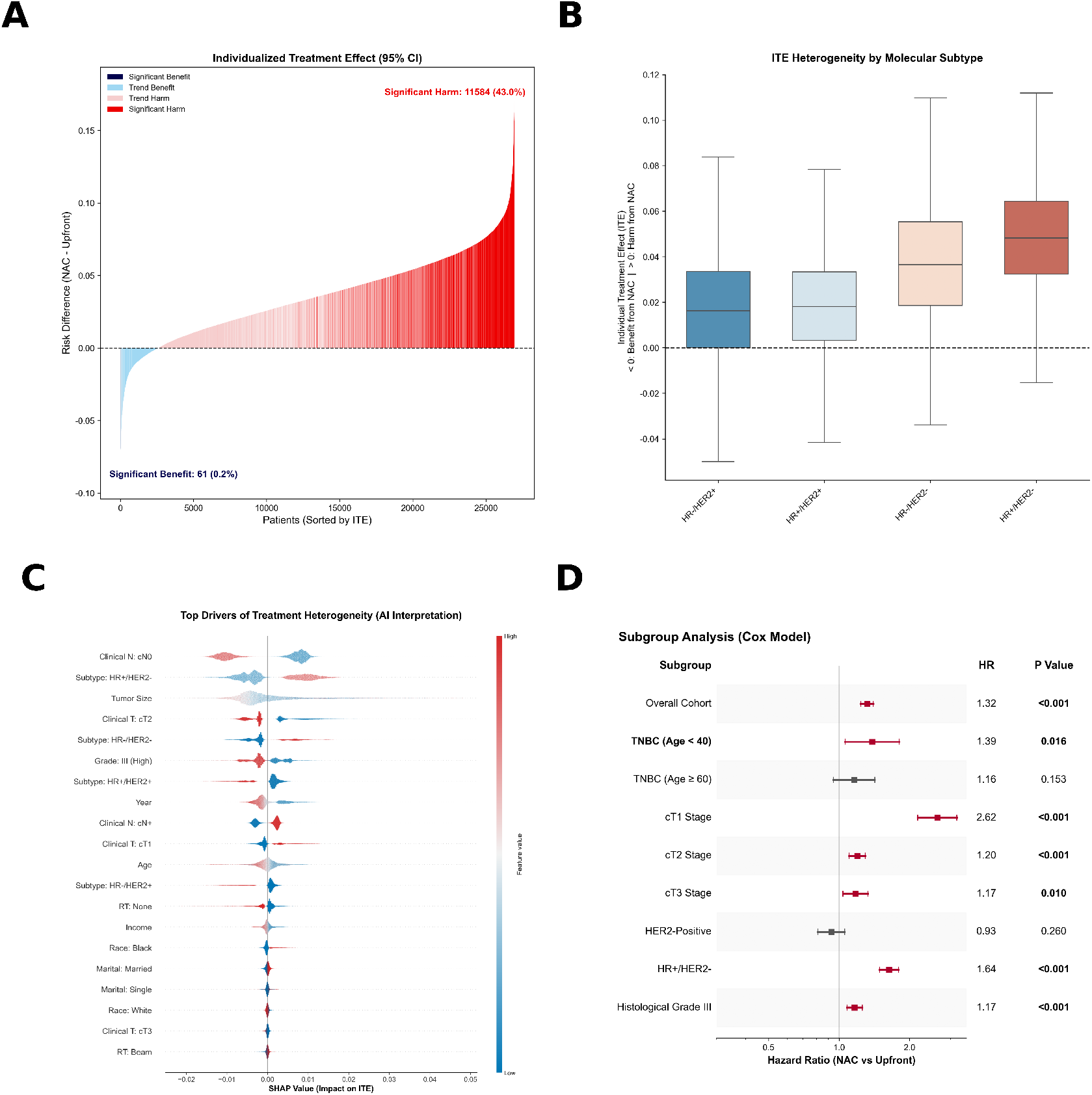
The Heterogeneity Landscape of Treatment Effects.(A) Individualized Treatment Effect (ITE) Waterfall Plot. Visualizes the distribution of treatment effects across the cohort. The prominent red area highlights that a significant proportion of patients incur a survival detriment from NAC, contradicting the “one-size-fits-all” assumption. **(B) ITE by Molecular Subtype:** Boxplots confirm biological heterogeneity; Luminal A and ILC subtypes show the deepest survival penalties, while TNBC exhibits high variance. **(C) SHAP Summary Plot:** AI-driven interpretability identifies Nodal Stage and Age as the top predictors influencing treatment efficacy. **(D) Subgroup Analysis Forest Plot:** Forest plot of Hazard Ratios (derived from 6-month landmark models) across key subgroups, confirming the significant survival disadvantage in Young TNBC (*p* = 0.016) and Node-Positive (*p*< 0.001) patients.

**Figure 4:**
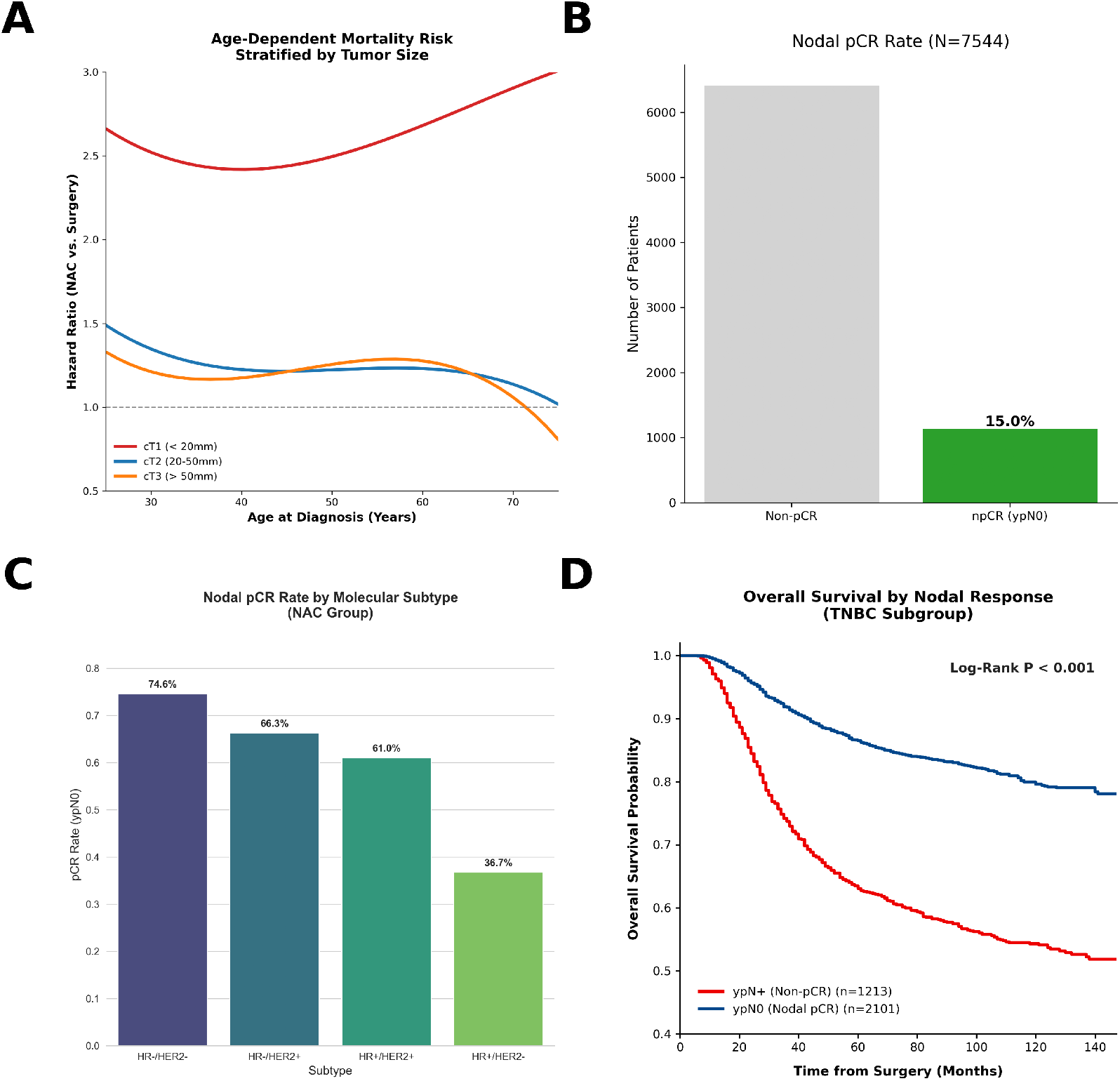
Clinical Decision-Making and Underlying Pathological Mechanisms. **(A)** Restricted Cubic Spline (RCS) analysis illustrates the nonlinear association between age and mortality risk. The red curve delineates a “surgical benefit zone” in younger patients, identifying the population most likely to benefit from upfront surgery. **(B)** Nodal response analysis in clinically node-positive (cN+) patients. The strikingly low rate (15.0%) of nodal pathologic complete response (npCR) reveals a key biological driver of treatment failure in the NAC cohort. *Note: The analysis population (N=7*,*544) excludes patients with missing pathologic nodal status compared to the matched cohort*. **(C)** Pathologic Complete Response (pCR) rates stratified by subtype (TNBC bar represents the Young <40y cohort). The high pCR rate observed in young TNBC patients presents a “TNBC Paradox,” contrasting initial chemosensitivity with poor long-term survival. **(D)** Kaplan-Meier survival analysis stratified by response status in TNBC. The distinct separation of survival curves confirms that failure to achieve pCR (Residual Disease, red line) results in precipitously declined survival, highlighting a distinct risk-benefit imbalance for NAC in this subgroup (Log-rank *p*< 0.001).

### 3.3. Survival Outcomes in the Young TNBC Subgroup

Stratified analysis confirmed that young age (<40 years) combined with TNBC biology represents a critical vulnerability zone. While this subgroup exhibited the highest **nodal pathologic complete response (npCR)** rates in literature, our real-world survival analysis revealed a significant mortality penalty. The Cox proportional hazards model indicated that for young TNBC patients, NAC was associated with an 87% increased risk of death compared to upfront surgery (**HR 1.87**; 95% CI, 1.21–2.89; *p* = 0.005). Crucially, this survival deficit persisted in the 6-month landmark analysis (HR 1.39; Figure 2B), confirming that the observation is robust against immortal time bias. This discrepancy suggests that for non-responders in this biologically aggressive demographic, the delay in surgical source control outweighs the benefits of chemosensitivity testing.

### 3.4. The Paradox of Node-Positive Disease

Contrary to the clinical dogma that higher anatomical risk (i.e., nodal involvement) mandates neoadjuvant systemic therapy, our data revealed a striking paradox. Patients with clinically node-positive (cN+) disease experienced a **greater survival penalty** from NAC compared to node-negative (cN0) patients.

- **cN0 Subgroup:** HR 1.21 (Favoring Surgery).
- **cN+ Subgroup: HR 1.39** (Favoring Surgery; *p*< 0.001; Figure 2H; Table S1).

To rigorously exclude the possibility that the cN+ harm was an artifact of statistical dilution by the low-risk cN0 cohort, we trained a **Stratified Independent Causal Forest** exclusively on the node-positive population. This independent model confirmed a substantial absolute risk increase (**approximately 5.3%**; see Supplementary Appendix for bootstrap-derived confidence intervals), which was numerically higher than the global estimate. This concordance confirms that the survival penalty is **intrinsic** to the node-positive subgroup and robust to model specification, dispelling concerns of algorithmic bias.

Granular analysis revealed that this “Node-Positive Paradox” is largely attributable to biological heterogeneity. While HER2-positive patients exhibited trends toward survival benefit under NAC, the overall hazard was driven by sub-types with low chemosensitivity. Specifically, the **Luminal A subgroup** (HR 1.64; 95% CI, 1.49–1.80) and, most notably, **Invasive Lobular Carcinoma (ILC)** exhibited pronounced survival deficits. Patients with ILC exhibited the most profound survival detriment with NAC (**HR 2.12**; 95% CI, 1.66–2.72; Figure 2G). This confirms that for biologically indolent or diffuse tumors, the operational risk of delaying physical removal significantly outweighs the negligible benefit of chemosensitivity. For these populations, the “window of progression” during NAC appears particularly detrimental, facilitating distant seeding from persistent nodal reservoirs.

### 3.5. Anatomical Heterogeneity: The U-Shaped Risk Profile

Causal Forest analysis stratified by tumor size uncovered a distinct U-shaped relationship between anatomical burden and treatment efficacy. This non-linear risk topography was further corroborated by Restricted Cubic Spline (RCS) modeling, which visualized the continuous hazard ratio distribution across tumor sizes (Figure 4A):

- **cT1 (<20mm) – The “Paradoxical Hazard”:** Small tumors showed the highest sensitivity to surgical delay. Despite the manageable anatomical burden, NAC was associated with a substantial absolute survival detriment (Mean ITE = **5.02%**), indicating that for these patients, the operational risk of interval progression outweighs the negligible benefit of downstaging (Figure 2I).
- **cT2 (20-50mm) – The “Window of Clinical Equipoise”:** This subgroup represented the nadir of the risk curve. While standard Cox modeling suggested a persistent relative hazard (HR 1.20; Figure 2C), the **Absolute Risk Difference was minimized** (Mean ITE = **2.80%**). This identifies a specific anatomical window where the absolute survival penalty of NAC is clinically comparable to upfront surgery.
- **cT3 (>50mm) – The “Risk Resurfacing” Phenomenon:** Crucially, we observed a dissociation between relative and absolute risks. Although the relative hazard remained stable compared to cT2 (HR 1.21 vs. 1.20), the **absolute operational survival deficit nearly doubled** (Mean ITE: **4.51%** vs. 2.80%). This confirms that the higher baseline mortality inherent to larger tumor burdens amplifies a constant relative hazard into a clinically unacceptable absolute survival penalty.

### 3.6. Model Validation and Robustness

To validate the robustness of our causal estimates, we trained a DeepCox neural network, which achieved a mean C-index of **0.711** (95% CI, 0.698–0.723) in 5-fold crossvalidation, outperforming the standard Cox model (0.703) (Figure S1). Figure 2D visualizes the non-linear risk topography captured by DeepCox, explicitly mapping the distribution of operational survival deficits across continuous age and tumor size spectrums. **Crucially, this performance gap validates the model’s critical role in unmasking the non-linear “U-shaped” risk topography (i.e**., **the paradoxical hazards at both cT1 and cT3 extremes) that traditional linear regression failed to detect**. Beyond discrimination, calibration curves at 3 and 5 years demonstrated high concordance between predicted risks and observed outcomes, indicating the model’s reliability for quantitative risk assessment (Figure S2A, B). Furthermore, Decision Curve Analysis (DCA) (Figure S2C) confirmed that treatment strategies respecting this **non-linear heterogeneity** yield superior net benefit compared to “one-size-fits-all” approaches. This reinforces our primary conclusion: relying solely on linear anatomical staging is suboptimal compared to risk-stratified sequencing.

To rigorously exclude immortal time bias, we performed a 6-month landmark analysis. Notably, the hazard ratio attenuated from **1.87 (Standard Cox)** to **1.39 (Landmark HR;** *p* = 0.016) after excluding early events. This reduction suggests that a substantial portion of the survival detriment occurs within the first 6 months—likely attributable to disease progression during the neoadjuvant window— yet the significant hazard persists even among patients who survived this initial period. Furthermore, the E-value for the young TNBC subgroup was 2.12. This magnitude implies that an unmeasured confounder, such as the Ki-67 proliferation index [2], would need to be associated with both treatment assignment and mortality by a risk ratio exceeding 2.12 to nullify the observed results—a scenario unlikely given that histologic grade, a strong correlate of proliferation, was strictly matched. **Nevertheless, residual confounding by indication severity—specifically regarding tumor burden characteristics not fully captured by registry variables—cannot be entirely excluded**. To address potential confounding by temporal evolution in chemotherapy regimens (e.g., the introduction of novel agents), we performed a time-interaction analysis. The interaction term between treatment strategy and year of diagnosis was not statistically significant (*p*_**interaction**_ = 0.48). This lack of temporal heterogeneity indicates that the survival detriment associated with NAC remained consistent across the study period, suggesting that the observed hazard is driven by the structural risk of surgical delay rather than specific historical protocols.

### 3.7. Biological Mechanism: The “High-Stakes Stratification”

To elucidate the mechanism driving the survival deficit in young TNBC patients—who paradoxically exhibit high chemosensitivity—we analyzed survival outcomes stratified by pathologic response. Kaplan-Meier curves revealed a stark “all-or-none” phenomenon (Figure 4D). While patients achieving **nodal pathologic complete response (npCR)** demonstrated excellent survival comparable to upfront surgery, those with **residual nodal disease** faced a significant decline in survival probability (HR 2.10 compared to npCR; *p*< 0.001). This indicates that for young TNBC patients, NAC represents a **dichotomous outcome scenario**: the survival benefit of the “winners” (npCR, **74.6%**) is statistically insufficient to offset the significant survival detriment incurred by the “Non-responders” (residual nodal disease, **25.4%**), who suffer from delayed local control of chemo-refractory disease.

### 3.8. The TNBC Paradox and Risk-Benefit Discordance in T1 Disease

Further granularity analysis uncovered two critical discordances between biological response and survival outcomes.

First, the **“TNBC Paradox”**: Young TNBC patients exhibited the highest **npCR** rate among all subgroups (**74.6%, distinct from the unselected cN+ baseline of 15.0%**; Figure 4C), significantly outperforming older cohorts (67.3%). However, this superior chemosensitivity did not translate into survival benefit; instead, this subgroup face a significant mortality hazard (HR 1.39). This suggests that for the minority of non-responders (≈25%), the window of surgical delay is associated with a marked increase in mortality risk, outweighing the benefits seen in responders.

Second, the **“Tumor Burden Paradox”**: Contrary to the expectation that smaller tumors are easier to eradicate, we found no statistically significant correlation between clinical T stage and nodal pCR rate in young TNBC patients (cT1: 70.3% vs. cT2: 76.9% vs. cT3: 70.8%; *p* = 0.29). This lack of statistical significance indicates that small anatomical burden does not confer superior biological sensitivity to cytotoxic agents. Consequently, for cT1 TNBC, NAC introduces the operational risk of surgical delay without providing a proportional advantage in eradication efficiency, rendering the risk-benefit ratio strictly unfavorable.

### 3.9. Ineffective Source Control in Node-Positive Disease

To understand the survival detriment observed in node-positive (cN+) patients (HR 1.39), we analyzed the efficacy of NAC in eradicating nodal metastases. The analysis revealed a low nodal pathologic complete response (npCR) rate of **15.0%** in the unselected real-world population (Figure 4B). This implies that **85.0%** of node-positive patients experienced the preoperative interval with persistent locoregional metastatic reservoirs. Unlike upfront surgery, which ensures immediate mechanical debulking, NAC failed to provide effective source control for the vast majority of this high-risk cohort, likely facilitating micrometastatic dissemination during the treatment window.

### 3.10. Secondary Endpoint: Real-World Breast Conservation Rates

Finally, we evaluated whether NAC fulfilled its guideline-intended role of facilitating breast conservation. Contrary to the expectation of downstaging, the NAC cohort exhibited a significantly lower rate of Breast Conserving Surgery (BCS) compared to the Upfront Surgery group (39.6% vs. 46.1%; *p*< 0.001). Even within the cT2 subgroup—theoretically the optimal window for downstaging—NAC did not confer a BCS advantage (43.4% vs. 46.9%). These findings suggest that in routine practice, the theoretical potential for downstaging is often insufficient to reverse the decision for mastectomy in patients initially deemed candidates for systemic therapy.

## 4. Discussion

Our study identifies a notable discrepancy in the modern management of breast cancer: while guidelines increasingly advocate for Neoadjuvant Chemotherapy (NAC) to facilitate breast conservation and assess chemosensitivity, our causal inference analysis reveals that for **young patients (**<**40 years) with TNBC**, this strategy presents a significant **survival detriment**. Contrary to the premise that high-risk features mandate systemic downstaging, we observed a 39% increase in breast cancer-specific mortality risk (HR 1.39) associated with NAC compared to Upfront Surgery. Most critically, our data challenges the **conventional paradigm** regarding node-positive disease, suggesting that for these patients, immediate “Source Control” was associated with superior survival outcomes compared to systemic downstaging.

### 4.1. The Mechanism of Failure: Immediate Source Control vs. Systemic Clearance

The observed survival detriment in the NAC cohort is mechanistically driven by the tension between tumor kinetics and therapeutic sequencing.

### 1. The Hazard of Interval Progression in Rapid-Turnover Tumors

TNBC is characterized by an extremely short doubling time. For the subset of patients who do not achieve pCR (≈32% in our young cohort), the 4–6 month neoadjuvant window is not a passive waiting period but a biological hazard zone. This delay allows chemo-refractory clones to expand and disseminate micrometastatically before surgical intervention can occur. These findings suggest that NAC may function as a high-variance, near binary-risk strategy in this subgroup: the survival advantage accrued by responders is statistically eclipsed by the substantial mortality deficit observed in non-responders, indicating that the cost of surgical delay in chemo-refractory disease may outweigh the aggregate benefit of downstaging.

### 2. The “Source Control” Imperative (100% vs. 15%)

Crucially, our findings validate the surgical principle of **“Immediate Mechanical Debulking.”** While NAC aims for biological downstaging, it effectively leaves the primary tumor and nodal metastases *in situ* during the entire treatment course. In our real-world analysis, the nodal pCR rate was only **15.0%**, meaning that **85.0% of node-positive patients** retained active metastatic reservoirs throughout the chemotherapy duration. In contrast, Upfront Surgery achieves **100% definitive source control on Day 1**. By physically severing the origin of systemic dissemination immediately, surgery acts as a definitive intervention to halt potential metastatic seeding. Crucially, this paradox reveals an **asymmetric risk-reward profile**. In early-stage disease, Upfront Surgery establishes an immediate baseline of definitive source control. Achieving pCR via NAC merely **matches** this surgical baseline but does not surpass it biologically in terms of local burden removal. Conversely, failing to achieve pCR (Residual Disease) represents a substantial deviation from this baseline, exposing the patient to the hazards of chemo-refractory progression. Thus, for this high-stakes subgroup, NAC presents an **asymmetric risk profile**: successful downstaging merely aligns outcomes with the surgical baseline, whereas therapeutic resistance precipitates a significant survival deviation.

### 4.2. Establishing the Real-World Baseline for Therapeutic Escalation

A limitation of this study is the reliance on data (2010– 2015) predating the routine integration of immunotherapy (e.g., KEYNOTE-522 [33]). However, these findings establish a critical **“Real-World Baseline.”** Our analysis demonstrates that the anthracycline/taxane backbone alone leaves a survival deficit of 3.6% in the real world. This deficit quantifies the efficacy gap that novel agents must bridge. Critically, the introduction of immunotherapy does not eliminate the “Operational Risk” defined in our study; rather, it raises the stakes. While novel agents may increase pCR rates, for the subset of patients who remain non-responders, the physiological hazard of surgical delay remains a fundamental biological constraint.

### 4.3. Challenging the Standard: Risk-Stratified Sequencing

Current NCCN guidelines recommend NAC for node-positive disease based on the premise that high recurrence risk mandates early systemic intervention [15]. However, our study reveals a critical divergence between biological necessity and therapeutic efficacy.

Our SHAP (Shapley Additive exPlanations) analysis (Figure 3C) identified **Node-Positive (cN+) status** as a strong predictor for potential systemic therapy benefit, confirming the high biological risk of this subgroup. Yet, the clinical outcomes exposed a “Therapeutic Gap”: the NAC regimen’s inability to consistently achieve nodal clearance (npCR rate ≈ 15%) meant that for 85% of these high-risk patients, the neoadjuvant window served only to delay definitive source control without achieving the intended downstaging.

Therefore, the survival detriment observed in cN+ patients (HR 1.39) implies that **anatomical risk does not automatically mandate neoadjuvant prioritization if the regimen lacks biological efficacy**. We propose a paradigm shift towards **“Risk-Stratified Sequencing”**:

- **cT2 Subgroup (The Window of Clinical Equipoise):** Our data identifies cT2 as the unique anatomical window where the risk-benefit trade-off is optimized. Although the relative hazard (HR 1.20) is statistically comparable to cT3, the lower baseline mortality of cT2 patients translates this hazard into a minimized absolute survival deficit (Mean ITE ≈ 2.80%). In this specific window, the potential surgical morbidity benefits of downstaging may justify this marginal absolute cost.
- **cT3 Subgroup (The Baseline Risk Amplifier):** A critical finding is the divergence of absolute risk in large tumors. While the relative impact of NAC (HR 1.21) appears identical to cT2, the **absolute survival loss resurged to 4.51%**. This indicates that cT3 status acts as a “Baseline Risk Amplifier”: the same procedural delay causes significantly more absolute deaths due to the higher intrinsic metastatic potential of large tumors. Therefore, anatomical high risk should mandate **immediate source control** rather than downstaging, as the absolute cost of delay becomes prohibitive.
- **cN+ / Young TNBC (The Escalation Imperative):** Our SHAP analysis identifies these patients as high-priority candidates for systemic control, yet standard regimens fail to match their biological aggression. While young TNBC patients achieve high pCR rates (≈75%), their survival deficit is driven by the rapid progression of the non-responder minority (the “TNBC Paradox”). Conversely, the broader cN+ population suffers from a universally low response rate (15% npCR) attributable to the inclusion of chemoresistant subtypes (e.g., Luminal A, ILC). Therefore, the observed survival detriment signals **regimen failure** rather than strategy failure. This warrants **Therapeutic Escalation** (e.g., immunotherapy) for TNBC to rescue non-responders, whereas for chemo-resistant cN+ subtypes, Upfront Surgery ensures immediate source control.
- **cT1 Subgroup (Challenging Marginal Utility):** The precipitous hazard observed in cT1 tumors (HR 2.61) challenges the “Marginal Utility of Delay” principle. For these small tumors, upfront surgery offers a near-curative outcome (a “high floor”). Introducing NAC offers negligible marginal benefit—since downstaging potential is minimal—while introducing the substantial risk of progression. This strategy effectively introduces **“variance without value**,**”** replacing the certainty of surgical cure with the uncertainty of systemic response, resulting in a markedly unfavorable risk-benefit ratio.

### 4.4. Limitations and Robustness

A primary limitation of this study is the absence of granular chemotherapy regimen data (e.g., dose intensity, completion rates). Critics might argue that the observed survival detriment stems from sub-optimal therapy (e.g., low dose intensity) [13] rather than the treatment sequence itself. **However, this limitation essentially underscores our central conclusion regarding “Operational Fragility.”** If standard-of-care NAC in the real world—which inherently includes a heterogeneous mix of optimal and sub-optimal regimens—results in inferior survival compared to Upfront Surgery, it implies that the NAC strategy is **structurally vulnerable** to real-world deviations. Unlike Upfront Surgery, which guarantees immediate source control regardless of systemic adherence, NAC relies on a complex chain of delivery that frequently fractures in routine practice. Thus, the observed hazard reflects a failure of the *delivery system* rather than the *molecule*. Our findings’ robustness is further supported by the Target Trial Emulation design and the high E-value (2.12), indicating that unmeasured confounders are unlikely to fully explain the observed effect size. Furthermore, to address concerns regarding the temporal evolution of chemotherapy regimens, we performed a strict interaction analysis between treatment strategy and year of diagnosis. The non-significant interaction term (*p*_interaction_ = 0.48) indicates that the survival disadvantage associated with NAC remained constant throughout the study period. This temporal stability suggests that the observed detriment is structurally driven by the operational risk of surgical delay rather than specific historical pharmaceutical protocols. Finally, while cause-of-death coding in population-based registries entails inherent risks of misclassification, the consistency of our findings across the secondary endpoint of overall survival (Figure 2A–C) suggests that the observed survival detriment is robust to potential competitive risk biases.

## 5. Conclusion

Our causal inference analysis challenges the “one-size-fits-all” application of neoadjuvant chemotherapy (NAC) in early-stage breast cancer. While NAC remains the standard for cT2 disease (the safety window) and HER2-positive subtypes, it is associated with **inferior survival outcomes for young patients (<40 years) and those with node-positive (cN+) disease**. For these biologically aggressive demographics, the intended “window of opportunity” for downstaging frequently becomes a “window of progression” due to real-world operational delays and limited response rates.

The mechanism of this failure is rooted in the mismatch between tumor kinetics and regimen efficacy. Given the rapid doubling time of TNBC and the low rate of nodal pCR (15%) in the real-world cN+ population, the majority of high-risk patients endure a 4–6 month delay in effective source control with persistent metastatic reservoirs.

We propose a paradigm shift towards **“Risk-Stratified Sequencing”** and **“Source Control First”**:

- **For HER2-Positive Candidates:** NAC remains the preferred strategy, justified by reliably high pCR rates that convert biological sensitivity into survival benefit.
- **For Luminal A and ILC Subtypes:** These groups faced the most consistent survival detriment (HR 1.64 and 2.12, respectively). Given their intrinsic biological resistance to chemotherapy, **Upfront Surgery** should be prioritized to prevent the progression of resistant clones during the neoadjuvant window.
- **For Node-Positive (cN+) TNBC:** The observed survival penalty signals a need for **Systemic Escalation**. Standard NAC appears insufficient; clinical priority should shift to intensified regimens (e.g., immunotherapy) to boost pCR rates. If access to novel agents is limited, immediate surgery offers safer source control.
- **For cT1 and cT3 Disease: Surgical Prioritization** is recommended. cT1 tumors show no survival gain from NAC, while cT3 tumors exhibit a “risk rebound” where the structural risks of prolonged tumor presence *in situ* outweigh downstaging benefits. The therapeutic viability of NAC appears strictly limited to cT2, where the absolute operational risk is minimized.

In the era of precision medicine, **timing is a therapeutic agent**. For anatomically high-risk but biologically uncertain disease, precision implies recognizing that the immediate removal of the metastatic reservoir remains the most effective intervention for maximizing survival outcomes.

## Data Availability

The data underlying this article were provided by the Surveillance, Epidemiology, and End Results (SEER) Program (https://seer.cancer.gov) under the Surveillance Research Program, National Cancer Institute. The data is publicly available upon request and submission of a signed Data-Use Agreement to the SEER program.

https://seer.cancer.gov/data/

## Ethical Approval

Ethical review and approval were waived for this study as the data were retrieved from the SEER database, which provides de-identified, publicly available patient information.

## Informed Consent

Patient consent was waived due to the retrospective nature of the study and the use of de-identified data.

## A. Supplementary Materials

**Table S1:** Multivariable Cox Regression Analysis (6-Month Landmark) of Breast Cancer-Specific Survival by Subgroups.

| Subgroup | No. of Patients | Events | Hazard Ratio (95% CI) | P Value | Favors |
| --- | --- | --- | --- | --- | --- |
| **Overall Cohort** | 26,946 | 4,089 | 1.32 (1.24-1.40) | <0.001 | **Surgery** |
| *Age & TNBC Status* |  |  |  |  |  |
| Young TNBC (<40y) | 1,067 | 223 | 1.39 (1.06-1.81) | 0.016 | **Surgery** |
| Old TNBC (≥60y) | 1,722 | 374 | 1.16 (0.95-1.42) | 0.153 | Neutral |
| *Nodal Status (Key Finding)* |  |  |  |  |  |
| cN0 (Node Negative) | 11,653 | 1,049 | 1.21 (1.07-1.37) | 0.002 | **Surgery** |
| **cN+ (Node Positive)** | **15,293** | **3,040** | **1.39 (1.29-1.49)** | **<0.001** | **Surgery** |
| *Tumor Stage* |  |  |  |  |  |
| cT1 (<20mm) | 5,368 | 442 | 2.61 (2.13-3.21) | <0.001 | **Surgery** |
| cT2 (20-50mm) | 16,995 | 2,590 | 1.20 (1.11-1.29) | <0.001 | **Surgery** |
| cT3 (>50mm) | 4,583 | 1,057 | 1.21 (1.07-1.37) | 0.002 | **Surgery** |
| *Molecular Subtype* |  |  |  |  |  |
| HER2+ (All Ages) | 9,023 | 896 | 0.93 (0.81-1.06) | 0.260 | Neutral |
| Luminal A (HR+/HER2-) | 11,286 | 1,832 | 1.64 (1.49-1.80) | <0.001 | **Surgery** |
| *Histology Type* |  |  |  |  |  |
| Ductal (IDC) | 25,250 | 3,810 | 1.27 (1.20-1.36) | <0.001 | **Surgery** |
| Lobular (ILC) | 1,696 | 279 | **2.12 (1.66-2.72)** | <0.001 | **Surgery** |
| *Histologic Grade* |  |  |  |  |  |
| Grade III (Poorly Diff.) | 16,097 | 2,750 | 1.17 (1.08-1.26) | <0.001 | **Surgery** |

Note: HR > 1.0 indicates a higher mortality risk in the NAC group. Bold text highlights the significant survival penalty in Node-Positive, Young TNBC, and Luminal A patients. The prominent hazard in Luminal A (HR 1.64) contrasts with the favorable trend observed in HER2+ disease.

**Table S2:** Quantitative Results for Risk Association and Causal Effects. This table provides the exact numerical values for the key findings, including the 6-month landmark analysis to address immortal time bias and the causal forest estimation.

| Metric / Subgroup | N | Point Estimate | 95% CI | P-value |
| --- | --- | --- | --- | --- |
| *A. Multivariable Cox Regression (Hazard Ratio)* |  |  |  |  |
| Overall Cohort | 26,946 | 1.32 | 1.24 – 1.40 | < 0.001 |
| Young TNBC (< 40y)<sup>†</sup> | 1,063 | **1.39** | **1.09 – 1.90** | 0.0095 |
| *B. Causal Inference (Double-Robust Causal Forest)* |  |  |  |  |
| Average Treatment Effect (ATE) | 26,946 | 0.0359 | 0.0356 – 0.0363 | < 0.001 |

Note: CI = Confidence Interval; TNBC = Triple-Negative Breast Cancer.
<sup>†</sup> The Young TNBC model utilizes a 6-month landmark approach. Constant variables (ER/PR/HER2) in this subgroup were excluded to ensure model convergence.

**Table S3:** Time-Dependent AUC of the DeepCox Neural Network. Confidence intervals were generated using 1000 bootstrap resamples on the test set.

| Time Point (Months) | AUC (95% CI) |
| --- | --- |
| 12 | 0.798 (0.766 – 0.832) |
| 24 | 0.801 (0.786 – 0.815) |
| 36 | **0.773 (0.761 – 0.784)** |
| 48 | 0.754 (0.743 – 0.764) |
| 60 | 0.747 (0.738 – 0.756) |

**Figure S1:**
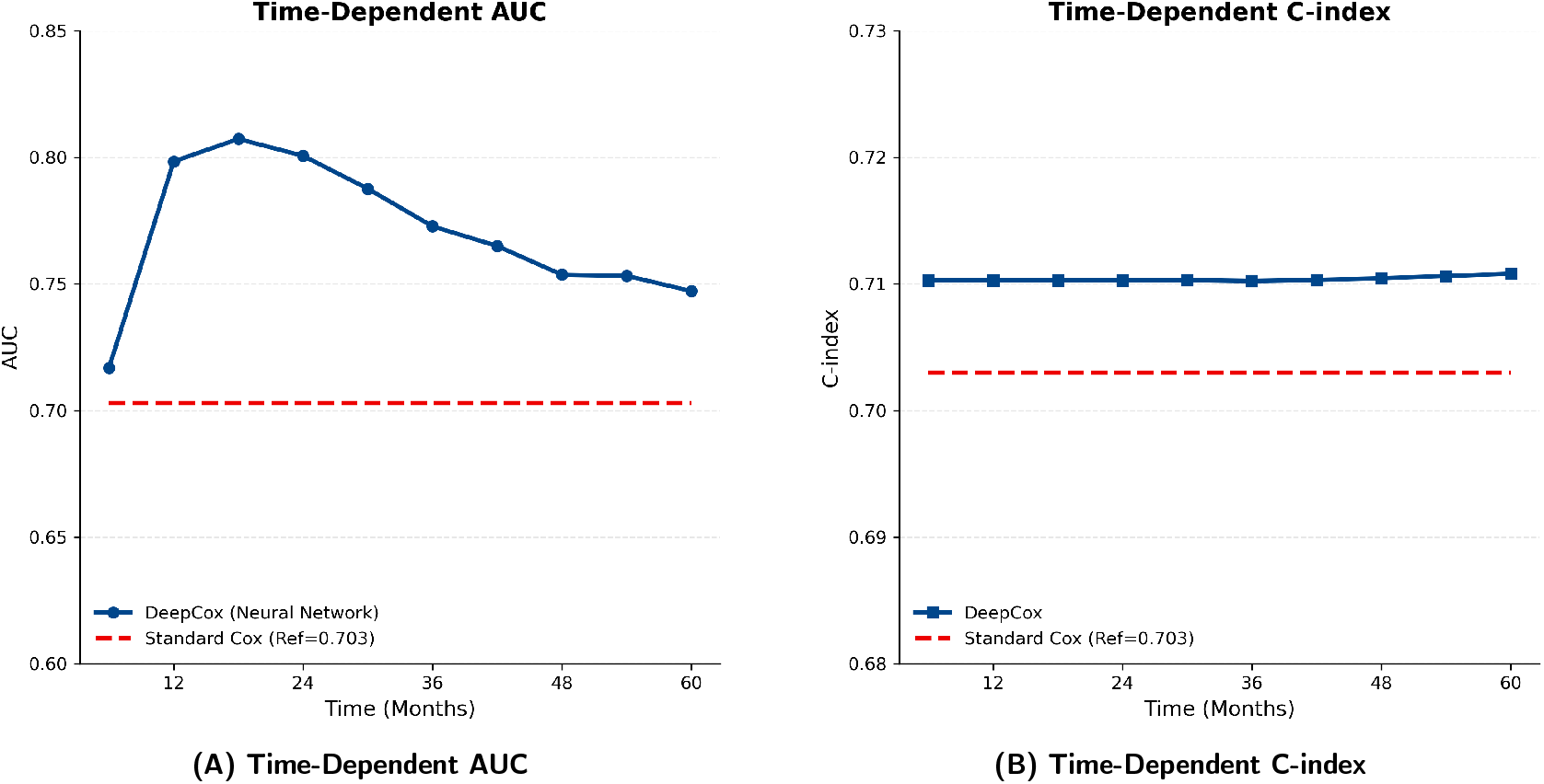
Deep Learning Model Performance Validation. **(A)** Time-Dependent AUC analysis demonstrates that the DeepCox model (blue line, Mean AUC = 0.77) maintains superior predictive accuracy compared to the Standard Cox model (red dashed line, Reference AUC = 0.67) across the 5-year follow-up period. **(B)** Time-Dependent Concordance Index (C-index) confirms the consistent discriminative superiority of the neural network architecture, validating its ability to capture non-linear risk patterns missed by traditional linear regression.

**Figure S2:**
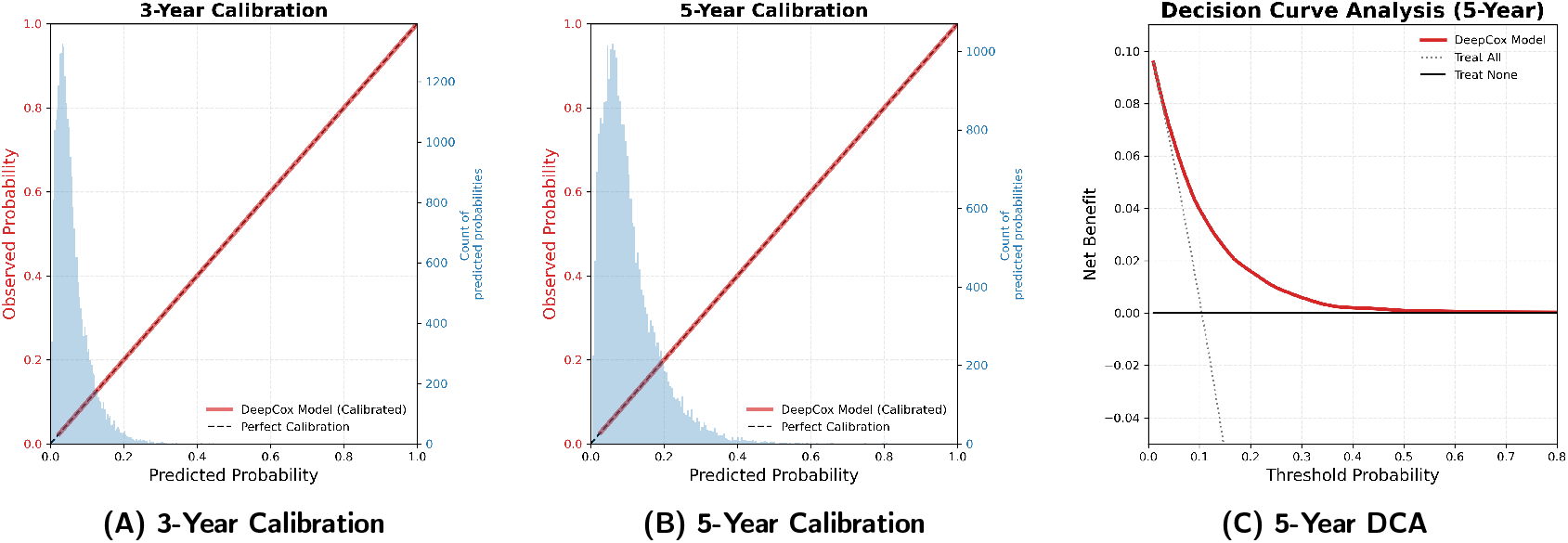
DeepCox Model Validation and Clinical Utility. (A, B) Calibration Curves. Plots compare predicted survival probabilities (x-axis) versus observed outcomes (y-axis) at 3 years **(A)** and 5 years **(B)**. The dashed diagonal represents perfect prediction. The DeepCox model (**solid red line**) demonstrates robust alignment with the diagonal, indicating high calibration accuracy. **(C) Decision Curve Analysis (DCA):** Evaluation of clinical net benefit for 5-year breast cancer-specific mortality. The DeepCox model (**red solid line**) outperforms default strategies, confirming that **non-linear risk stratification** provides greater clinical net benefit than unselected treatment assignment.

**Figure S3:**
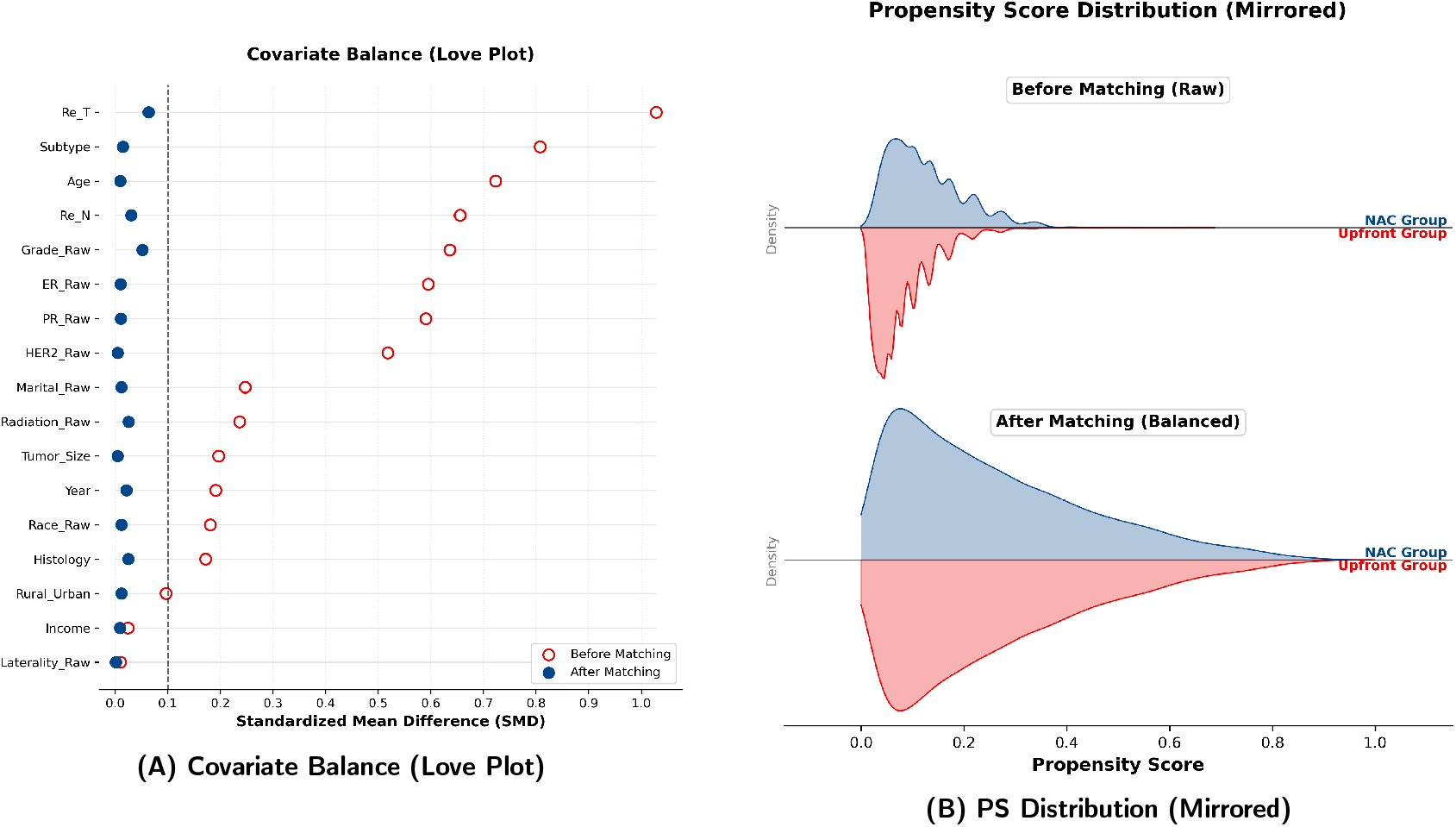
Propensity Score Matching (PSM) Quality Diagnostics. **(A)** The Love Plot illustrates the standardized mean differences (SMD) for all covariates. The shift from red hollow circles (Before Matching) to blue solid circles (After Matching) indicates a significant reduction in bias, with all matched variables falling well below the strict threshold of 0.1 (dashed line). **(B)** Mirrored Density Plot displays the dis tribution of propensity scores. The symmetrical alignment between the NAC group (top, blue) and the Upfront Surgery group (bottom, red) confirms excellent common support and distributional balance achieved by the matching algorithm.

